# The cost of myopic pandemic response

**DOI:** 10.1101/2024.02.19.24303020

**Authors:** Sarah Nowak, Pedro Nascimento de Lima, Raffaele Vardavas

## Abstract

Prior to the availability of COVID-19 vaccines, non-pharmaceutical interventions (NPIs) served as a primary strategy to mitigate the spread of the disease. However, the efficiency of these interventions relies on understanding and incorporating human behavior into infectious disease models. This study addresses the need for models that better account for the influence of temporal discounting on behavioral dynamics to enhance forecasting accuracy and develop robust mitigation strategies. Our previous research introduced Known Time Horizon (KTH) policies, optimizing social distancing measures based on a central planner’s rational assessment of the pandemic’s time frame and associated costs. In this paper, we contrast the KTH policy with a model reflecting myopic decision-making, an extreme form of temporal discounting that emphasizes short-term outcomes over long-term consequences. By comparing the expected social distancing behavior under myopic decision-making with the optimal policy derived from KTH approaches, we elucidate the impact of temporal bias on social distancing practices and assess its implications for infection dynamics and associated costs. We find that myopic policy always results in greater total costs throughout an epidemic compared to a KTH policy. However, each cost component – the costs of infection and social distancing – derived from a myopic strategy may be either larger or smaller than the component costs for a strategy developed using a full optimization model, depending on the specific parameters involved as myopic decision-makers seek to delay both costs of social distancing and infection.

## 1 Introduction

In the early phases of the COVID-19 pandemic, before vaccines became available, public health officials relied on non-pharmaceutical interventions (NPIs) to slow the spread of the disease.^1–3^ While infectious disease modeling was used to inform public health response efforts, it has become clear that models need to better incorporate behavior to improve forecasting^4–6^ and the development of optimal and robust mitigation strategies.^7–9^ In addition, there is a mismatch between the appropriate time-frame to be considered in decision-making (i.e., timeframe of the pandemic which is months to years) and the time-frame over which predictive pandemic forecasting models are accurate (a few weeks). This mismatch creates a situation in which policymakers do not have access to estimates of the effects of their policy choices over the relevant time frame of the impact of their decisions.

In previous work, we formulated a parsimonious model to calculate optimal social distancing policies.^10^ Our analysis identified optimal NPI trajectories based on a central planner’s assessment of the pandemic’s time horizon and the costs of social distancing relative to infection-related costs. Employing a deductive reasoning approach that assumes a society governed by a rational central planner, we modeled behavior using a deterministic susceptible-infectious-recovered (SIR) model and calculus of variations-based optimization approach, which resulted in a boundary value problem (BVP). We found two qualitatively different classes of policy responses, which, for simplicity, we labeled: **suppression**, in which transmission levels were kept very low, and **mitigation**, in which herd immunity was reached. Our findings showed that the central planner’s decision between these strategies relied on two key factors: the cost of social distancing relative to the cost of infection and the pandemic time horizon, which is determined by a central planner’s expectations regarding when effective pharmaceutical interventions like vaccines would become accessible. Here, we will refer to the policies derived in our prior work as Known Time Horizon (KTH) policies.

In this paper, we compare the KTH policy with the expected social distancing behavior under the assumption of *myopic* decision-making - i.e, decision-making that only weighs short-term outcomes. At each moment, a central planner weighs the costs of social distancing in the next infinitesimal time step against the costs of infection in the next infinitesimal time step. It is well-documented that individuals tend to give greater consideration to near-term losses and gains than to long-term outcomes, a phenomenon known as present bias or temporal discounting.^11,12^ Myopic decision-making represents an extreme form of temporal discounting where only immediate consequences are considered and the distant future is ignored. While myopic decision-making may be an extreme assumption, such a model serves as a valuable tool to explore the impact of short-term bias on social distancing behavior and to gauge how social distancing, infection dynamics, and costs compare to a scenario in which an optimal policy is pursued.

## 2 Methods

### 2.1 Dimensionless SIR model

There are three key parameters in the non-dimensional specification of our model: *c, τ*_final_, *R*_0_. The first parameter is *c*, which quantifies how individuals perceive the distress or burden of practicing social distancing relative to the costs of getting infected. A small *c* close to 0.1 indicates a preference for health and safety, while a large *c* indicates a preference for social and economic activities even in the face of potential health risks. Next, we consider *τ*_final_, representing the time horizon. This parameter marks the point at which a central planner envisions the availability of pharmaceutical interventions that make social distancing unnecessary. *τ*_final_ is expressed in terms of the non-dimensional time, *τ*, where each unit corresponds to *γ*^*−*1^, the average time to recovery. The last parameter is *R*_0_ - the initial reproduction number describing the baseline transmission rate. In the early stages of the epidemic, when few individuals are infected, *R*_0_ is the average number of new infections generated by each existing infection, assuming no preventive measures such as social distancing. In our model, we solve for *R*_*D*_, the dynamic reproduction number, which is lower than *R*_0_ when social distancing policies are in place.

For this analysis, we solve a system of differential equations characterizing the distribution of individuals across susceptible (*s*), infected (*i*), and recovered (*r*) states within the SIR model:

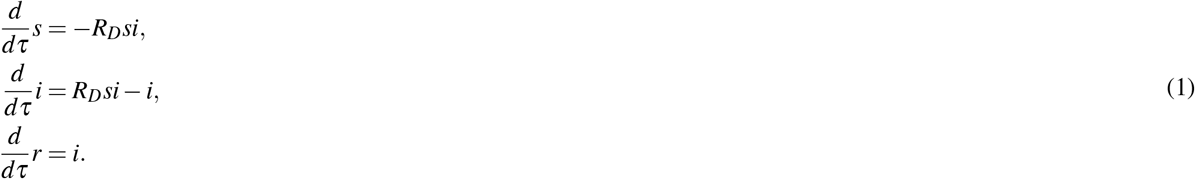

Here, *R*_*D*_ represents a dynamic reproduction number, which is reduced as individuals reduce infectious mixing (either by policy or behavioral change).

### 2.2 Myopic and Optimal Pandemic Response Policies

Assuming myopic behavior, *R*_*D*_ is chosen to minimize the instantaneous cost of social distancing plus the instantaneous cost of new infections. In contrast, the optimal decision weighs all costs and benefits over the *τ*_final_ time horizon. The instantaneous total cost *h* of the pandemic is expressed as:

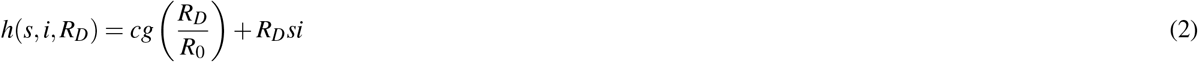

The first term, *cg*(*R*_*D*_*/R*_0_), denotes the cost of social distancing per unit time, where *g*(*x*) = *−* ln(*x*) + *x −* 1. This function ensures a decreasing cost as *R*_*D*_ increases, with a minimum at 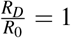. The second term in equation 2 is the cost of new infections per unit time, equivalent to the rate of new infections in the dimensionless form.

To find a myopic solution minimizing the instantaneous cost, *R*_*D*_ is chosen such that:

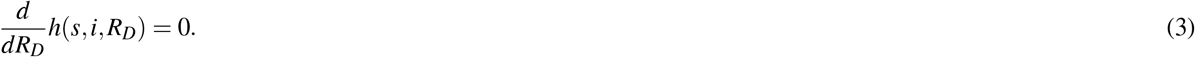

This leads to:

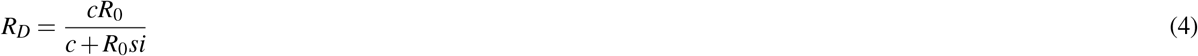

Equation 1 combined with 4 defines the dynamics of the myopic solution. The estimated *R*_0_ for the 2009 influenza pandemic was 1.5, and for COVID-19, it ranges from 1.8 to 3.6 with an estimated value of 2.5^13–15^.Further details on the non-dimensional model and the solution approach for the optimal solution can be found in the Methods Appendix.

We obtain the time-varying *R*_*D*_ for both the myopic and KTH policies, and compare key outcomes, including the total epidemic since, the total cost of the pandemic. We present the “cost” (also known as regret) of the myopic policy compared to the KTH policy.

## Results

In our previous research, we modeled a rational central planner who would impose an optimal social distancing policy given assumptions about *τ*_final_, *R*_0_, and *c*. We described two qualitatively distinct classes time-varying social distancing strategies that emerged from the KTH model. One strategy, a “suppression” strategy, involved an initial “lockdown” with a low *R*_*D*_ that gradually increased over time, keeping infections low. The other strategy, a “mitigation” strategy, featured little initial social distancing if infections were low, with increased distancing as infections climbed, and *R*_*D*_ decreasing to its lowest point near the infection peak. In mitigation strategies, herd immunity is achieved and the population can fully re-open by the end of the time horizon. When the economic, social, and other costs associated with social distancing measures are high, policymakers may be inclined to explore strategies that mitigate these costs. This could involve allowing more normal economic and social activities to continue, even if it comes at the expense of a potentially higher rate of infections. Moreover, if the costs associated with a suppression strategy are deemed unsustainable over an extended period, policymakers might opt for a mitigation strategy that allows for more normalcy, even if it means accepting a certain level of infection. Hence, when the costs of social distancing are high relative to infection (i.e., a high value for *c*), disease transmissibility is challenging to control (i.e., a high value for *R*_0_), and the epidemic time horizon is long (i.e., a high value for *τ*_final_), a rational policymaker will pursue a mitigation strategy prioritizing livelihoods over saving lives. However, our findings indicate a sharp, non-linear transition from mitigation to suppression when the relative social distancing cost, transmissibility, or time horizon, or a combination of these, falls below a critical threshold. Our findings emphasized the importance of public messaging and provided a mathematically consistent explanation for limited popularity and adherence to NPIs in specific regions.

Figure 1 compares the social distancing strategies defined by *R*_*D*_ under the KTH policy (solid line) from our prior work^10^ and a myopic policy (dashed line). Figure 1a shows the social distancing dynamics, which are shown as the dynamic reproduction number normalized by the baseline reproduction number, *R*_*D*_*/R*_0_. These dynamics are shown for different values of the relative cost of social distancing, *c*, and different values of the baseline reproduction number, *R*_0_. We find that qualitatively, the social distancing dynamics resemble the distancing dynamics for the mitigation strategy from our prior work. In the myopic solution, distancing does not begin until infection rates begin to climb, similar to the mitigation dynamics we found previously. In instances of low *c* and *R*_0_, the KTH model favors a suppression strategy. Conversely, the myopic policy leans towards a mitigation approach with considerably less social distancing (i.e., a dynamic with higher values of *R*_*D*_*/R*_0_) than the KTH policy. For higher *c* and *R*_0_ values, both the KTH policy and the myopic policy favor a mitigation strategy, but with notable distinctions. Although qualitatively similar, the myopic solution’s *R*_*D*_*/R*_0_ dynamics lead to a lower minimum valley of *R*_*D*_*/R*_0_, with social distancing persisting longer compared to the mitigation KTH policy. The myopic policy exhibits a more cautious mitigation strategy where, after reaching significant infection rates, social distancing is more stringent, and the relaxation of social distancing policies occurs at a slower rate than in the KTH policy.

**Figure 1.**
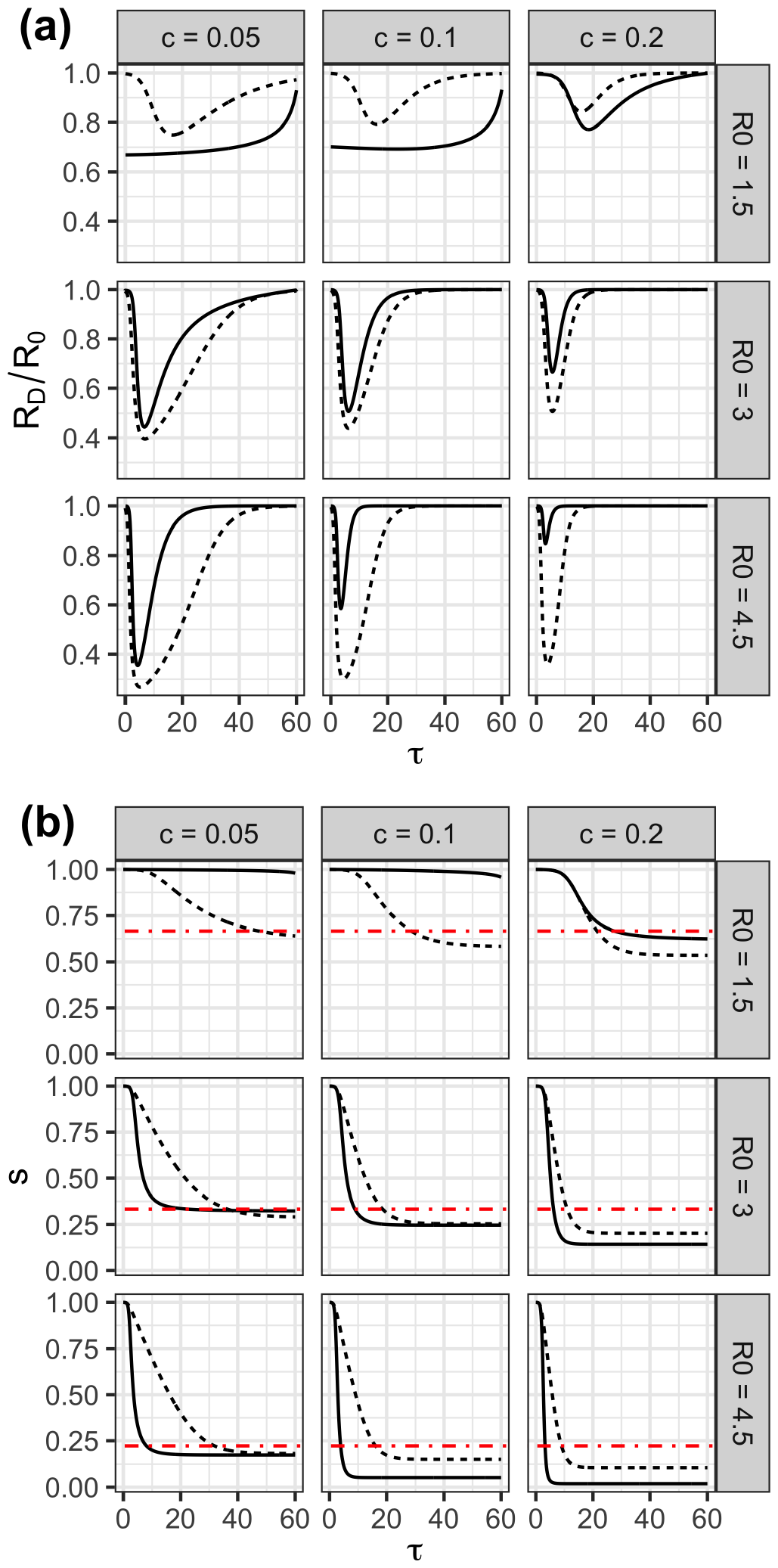
Comparison of distancing and infection dynamics under the KTH policy (solid line) or under the myopic strategy (dashed line). The red line shows the maximum number of susceptibles that can remain in the population after herd immunity is achieved. The relative cost of social distancing, *c*, is varied over the panel columns and the reproduction number, *R*_0_, is varied over the rows. (a) *R*_*D*_*/R*_0_, the dynamic reproduction number over time. (b) The proportion of susceptible individuals in the population over time. In all panels, the initial proportion of the population infected is *i*_0_ = 0.0001 and *τ*_final_ = 60.

Figure 1b depicts the dynamics of *s*(*τ*), representing the proportion of the population in the susceptible state, over time for both myopic and KTH policies. When the KTH policy is a suppression strategy (at low values of *c* and *R*_0_), the susceptible population remains close to 1 throughout the period, indicating a minimal number of cumulative infected cases. In these scenarios, the myopic policy exhibits a quicker drop in the number of individuals in the susceptible state compared to the KTH policy, resulting in a substantial proportion of the population ultimately being infected. By the fixed final epidemic time *τ*_final_, the myopic policy consistently yields a higher number of cumulative infected cases than suppression strategies resulting from the KTH policy.

As *c* increases, while still at low values of *R*_0_, the KTH policy transitions from suppression to a mitigation strategy, leading to a significant increase in cumulative infected cases. At low values of *R*_0_, the myopic policy continues to result in a larger number of cumulative infected cases compared to the KTH policy. For high *c* and *R*_0_ values, the situation reverses, with the KTH policy having a larger number of cumulative infected cases than the myopic policy. This suggests a crossover point between the KTH policy or the myopic policy having the most cumulative infected cases by *τ*_final_. The crossover is evident, particularly around the case where *c* = 0.05 and *R*_0_ = 3 (leftmost column, middle row of Figure 1b).

This case is intriguing because, despite *R*_*D*_ consistently being lower in the myopic case throughout the dynamics, more individuals are ultimately infected in the myopic solution, resulting in a lower final *s*. This apparent paradox is clarified by considering the timing of herd immunity and the effective reproductive number given by *R*_*t*_ = *R*_*D*_*s*. The KTH policy, with a faster infection rate, more quickly reaches the point of herd immunity, after which very few people continue to be infected. This is akin to the scenario where a vaccine is rapidly distributed, reaching herd immunity when a proportion equal to 1 *−* 1*/R*_0_ of the population is vaccinated, leaving 1*/R*_0_ of the remaining population susceptible. As this condition is met early in the dynamics within the KTH policy, its effective reproductive number (*R*_*t*_) decreases rapidly and stays consistently lower than the effective reproductive number of the myopic solution for the rest of the duration (see Figure S1 in the supplemental information). Consequently, a larger proportion of individuals, more than 1 *−* 1*/R*_0_, get infected by the final time *τ*_final_ in the myopic solution. Figure 2a shows the infection cost, which is equivalent to the final epidemic size, as a function of *c*, the relative cost of social distancing, *τ*_final_, the time horizon, and *R*_0_, the reproduction number. The total infection cost is shown for both KTH and myopic policies. As we found in our prior work^10^, the KTH policy results in a final epidemic size that displays a discontinuity as a function of *c* and *τ*_final_. Low values of the final epidemic size are the result of suppression strategies and moderate to high final epidemic sizes are the result of mitigation strategies. In contrast, final epidemic sizes resulting from a myopic strategy change continuously as a function of *τ*_final_ and *c*, with larger epidemic sizes resulting from longer time horizons (*τ*_final_), larger relative cost of social distancing (*c*) and higher reproduction numbers (*R*_0_). Figure 2b shows the differences between the final epidemic size for the myopic and KTH policies. When the KTH model leads to a suppression strategy, the final epidemic size is always larger for the corresponding myopic policy compared to KTH policy. For parameters where the KTH policy is a mitigation strategy, the myopic policy tends to result in a slightly smaller final epidemic size compared to the KTH policy for larger values of *c* and a slightly larger final epidemic size for smaller values of *c*.

**Figure 2.**
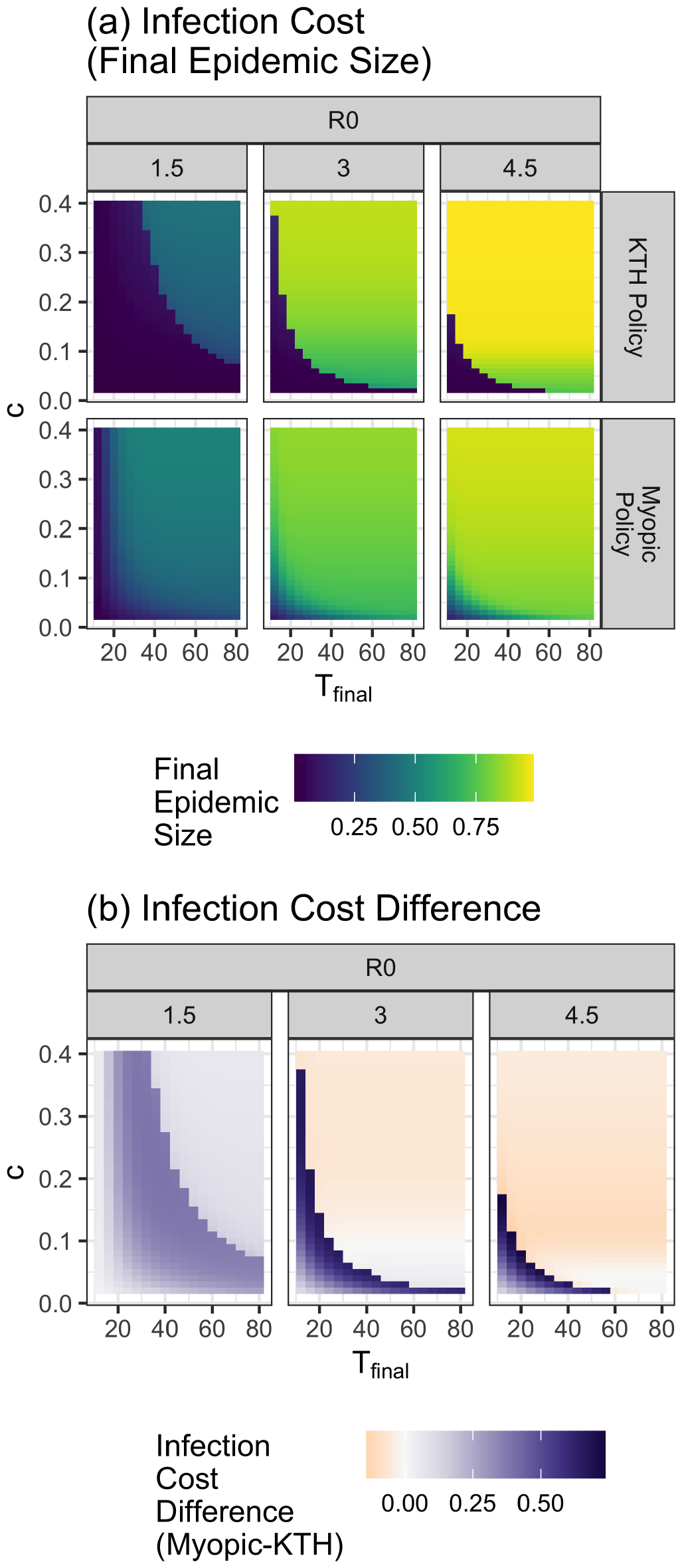
(a) Infection cost for the KTH and myopic policies. The infection cost is equal to the final epidemic size in our framework. (b) The differences between the myopic policy infection cost and the KTH policy infection costs are shown. For all runs, *i*_0_ = 0.0001. Values of *R*_0_, *τ*_final_, and *c* are shown in the figure. A cost of 1 is equivalent to the cost of the entire population becoming infected.

Figure 3a shows the social distancing costs for the KTH and myopic policies. In the KTH policy, at low values of *τ*_final_ and *c*, social distancing costs are low. They increase with increasing time horizon (*τ*_final_) and relative cost of social distancing (*c*) as long as the optimization results in a suppression strategy. Social distancing costs are greatest just before an increase in *c* or *τ*_final_ would result in a change from a suppression to a mitigation strategy. In contrast, the social distancing costs for the myopic strategies increase continuously and monotonically as a function of increasing *τ*_final_ and *c*. Figure 3b shows the difference in social distancing cost for the myopic and KTH policies. For *R*_0_ = 1.5, myopic social distancing costs are always lower than distancing costs for the KTH policy. For *R*_0_ = 3 and *R*_0_ = 4.5, social distancing costs are lower in the myopic policy when the KTH policy results in a suppression strategy and greater in the myopic case when the KTH policy results in a mitigation strategy.

**Figure 3.**
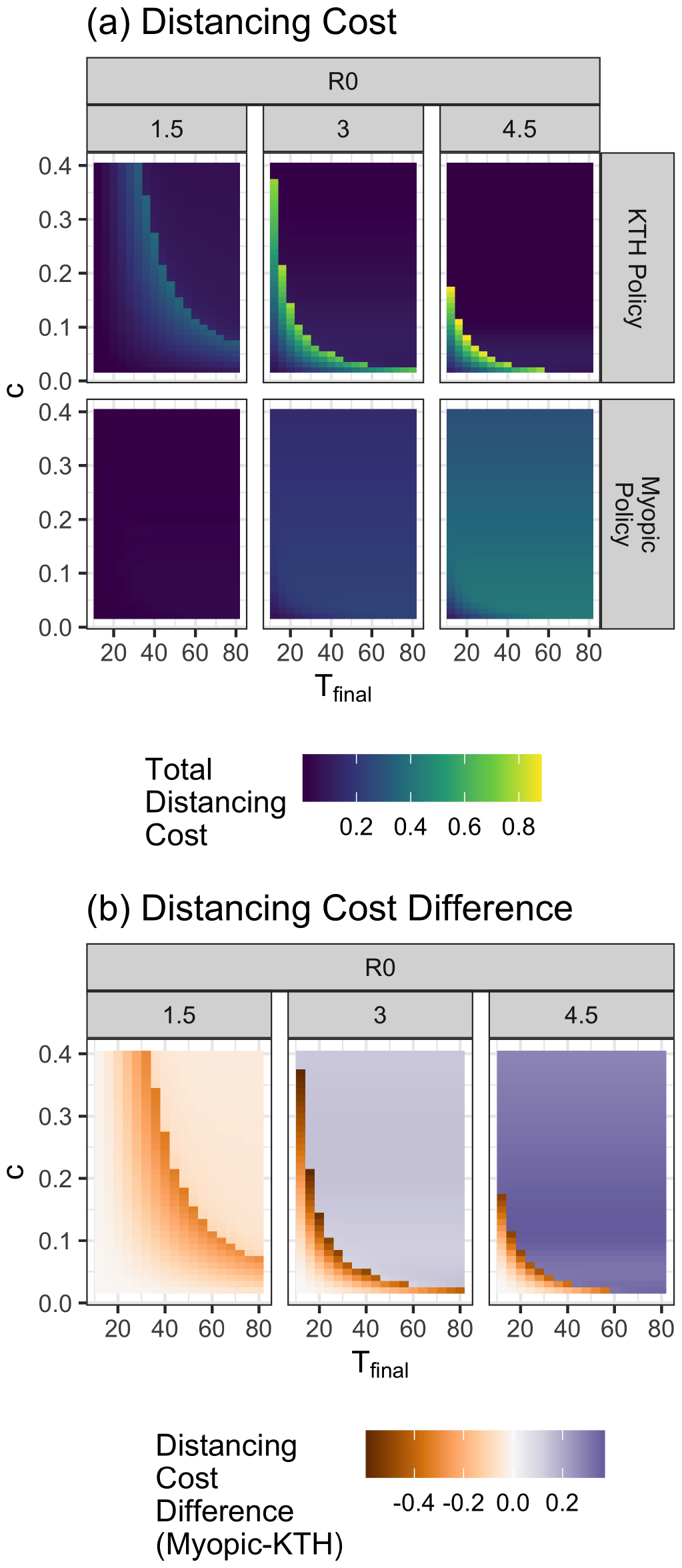
(a) This figure shows the distancing cost for both the KTH and myopic policies. (b) The differences between the myopic distancing cost and the KTH distancing cost are shown. Values of *τ*_final_, *c* and *R*_0_ are shown in the figure. *i*_0_ = 0.0001. A cost of 1 is equivalent to the cost of the entire population becoming infected.

Figure 4a shows the total cost of the KTH policy and from a myopic policy. Total costs for both policies increase continuously and monotonically with increasing *τ*_final_ and *c*. Total costs for the KTH policy are never greater than 1 (the cost of the entire population becoming infected), while costs for the myopic policy are close to 1.25 for *R*_0_ = 4.5 and larger values of *τ*_*rmfinal*_ and *c*. Such strategies are clearly inefficient as an uncontrolled epidemic would require no social distancing and result in less than the full population becoming infected. Figure 4b shows the difference in total cost for the myopic and KTH policies. The cost of the myopic policy is always greater than the cost of the KTH policy, as we expect. The myopic policies are most inefficient (most exceed the cost of the KTH policy) near the boundary where the KTH policy transitions from a suppression to a mitigation strategy.

**Figure 4.**
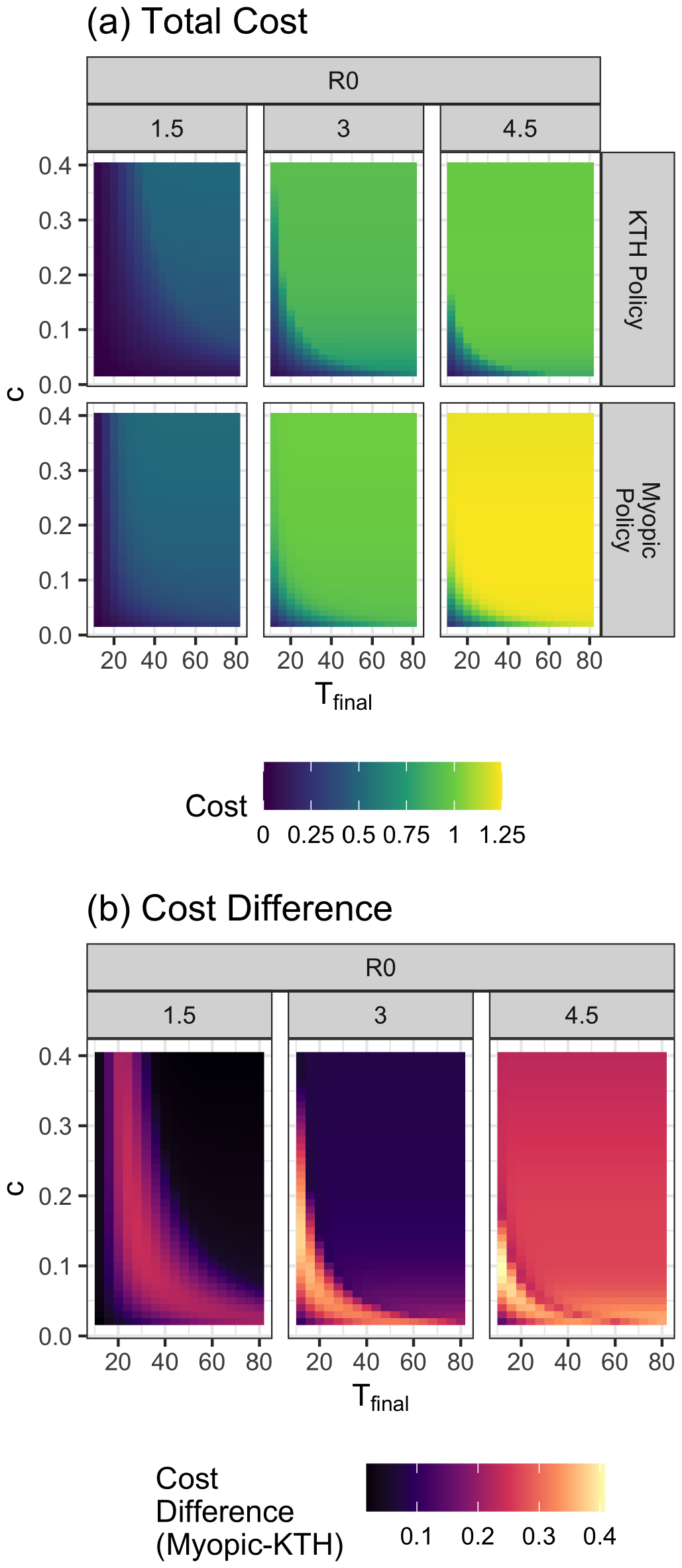
(a) Total costs for the full optimization and myopic solutions. (b) Difference in total costs for solutions resulting from the myopic solutions and full optimization. Values of *τ*_final_, *c* and *R*_0_ are shown in the figure. *i*_0_ = 0.0001. A cost of 1 is equivalent to the cost of the entire population becoming infected.

## Discussion

In prior work, we developed a model to find optimal social distancing strategies given the relative cost of social distancing compared to the cost of infection and the time horizon until pharmaceutical interventions become available.^10^ In that work, we found two qualitatively distinct types of optimal social distancing strategies: a suppression strategy in which infections are kept as low as possible, and a mitigation strategy in which herd immunity is reached. In this paper, we find that if individuals follow a myopic social distancing strategy, the distancing dynamics are qualitatively similar to the mitigation strategy found in our prior work. Total costs of social distancing plus infections are higher with a myopic social distancing strategy than with a strategy developed by a central planner through a full optimization, as expected.

If the full optimization model results in a **suppression strategy**, the myopic solution will result in a larger epidemic size and have a lower cost of social distancing compared to the full optimization. If the full optimization results in a **mitigation strategy**, the myopic solution sometimes results in a somewhat smaller epidemic size, generally for larger values of the baseline reproduction number, *R*_0_, and for larger values of *c*, the relative cost of social distancing compared to the cost of infection. When the full optimization leads to a mitigation strategy, the myopic solution results in lower social distancing costs compared to the full optimization model when *R*_0_ = 1.5, but leads to a higher cost of social distancing when *R*_0_ = 3 and *R*_0_ = 4.5.

Myopic decision-making is an extreme form of present bias, also known as temporal discounting, and our results therefore shed some light on how present bias may affect decision-making in a pandemic. One might naively assume that present bias leads individuals and policymakers to delay social distancing, instead seeking the instant gratification of being able to gather.

However, our results show that engaging in extreme present bias – myopic decision making – sometimes leads to individuals engaging in *more* distancing compared to a strategy developed by a central planner optimizing total costs over a defined time horizon. It is important to recognize that present bias leads individuals to attempt to delay not only the costs of social distancing, but also to delay the costs of infection. When the full optimization leads to herd immunity (using a mitigation strategy), the myopic solution usually also achieves herd immunity but takes longer to reach that point.

We note that our full optimization model does not consider many factors that would warrant slowing infections, even if reaching herd immunity is inevitable. For example, we do not consider any capacity limitation of the healthcare system which could result in higher per-individual infection costs if many individuals are infected simultaneously. Alternatively, it might be reasonable to consider that per-individual costs of infection decrease over time as the standard of care improves and treatments are introduced. Such effects could lead to optimal solutions that slow transmission earlier in the pandemic more than our current full optimization model, much as the myopic solutions do.

In a boundary value optimization, we explored the strategy - suppression or mitigation - that a rational policymaker would adopt if they knew the pandemic time horizon.^10^ In reality, the policymaker would not possess precise knowledge of the epidemic time horizon. Instead, they may have a bell-shaped distribution around their expected epidemic time horizon, reflecting their uncertainty and prior beliefs. However, other types of prior beliefs are possible^16^. A pessimistic planner might assume the remaining time until the end of the pandemic is proportional to the amount of time already passed. A more common prior is the uniform, memory-less prior. Myopic decision making is an extreme form of such decision-making in which the general public and policymakers make decisions based on the current epidemiological state without considering how long the pandemic has lasted or projecting into the future. Moreover, policymakers may adopt a strategy but be subject to a cost constraint (i.e., social distancing costs must not surpass some threshold parameter beyond which bearing the costs of distancing becomes unbearable). The population in society may have diverse expectations and priors, which may differ from those of policymakers. Some individuals in society may have a prior more aligned with a myopic, memoryless approach. Understanding the disparities between the policymakers’ prior and the societal population distribution priors may illuminate tensions between policymakers’ preferences and societal expectations. When policymakers and the public have different expectations, policymakers may need to incorporate societal expectations in their planning to improve compliance. Our paper offers initial insights into this critical question when society engages in myopic decision-making and a central planner optimizes over fixed time frame.

## 3 Conclusion

Individuals and policymakers may all use some level of present bias, also known as temporal discounting, when making decisions or setting policies about social distancing during a pandemic. Myopic decision-making, in which only very near-term costs and benefits of a policy are considered, is an extreme form of present bias, which yields insight into the consequences of present bias when making decisions about social distancing in the context of an epidemic. We find that myopic policy always results in greater total costs throughout an epidemic compared to a KTH policy. However, each cost component – the costs of infection and social distancing – derived from a myopic strategy may be either larger or smaller than the component costs for a strategy developed using a full optimization model, depending on the specific parameters involved.

## Data Availability

All code used is available at: https://github.com/pedroliman/covid19-bpv/tree/myopic

## Availability of Data and Materials

Code and data generated for the current study are available at https://github.com/pedroliman/covid19-bpv/tree/myopic.

## Acknowledgements

We wish to thank the National Cancer Institute (R21CA157571), and the National Institute of Allergies and Infectious Diseases (R01AI118705 & R01AI160240) for providing support in projects that led to preliminary work and ideas that motivated this project. Dr. Nowak acknowledges support from the Blodwen S. Huber Early Career Green and Gold Professor in Pathology and Laboratory Medicine at The Robert Larner, M.D. College of Medicine as well as support from The National Institute of General Medical Sciences 3P20GM125498-04S1 and 2P20GM125498P-06.

