## Supplement for "The cost of myopic pandemic response"

February 19, 2024

### 1 Methods

Note: The set up for this model is similar to that in [Nowak et al.(2023)Nowak, Nascimento de Lima, and Vardavas]
As a result, this supplement has some material in common with the supplement of that paper.

#### 1.1 The SIR Model

We use a standard Susceptible, Infectious, Recovered (SIR) model of disease transmission. The standard SIR model describes disease transmission in a well-mixed system in which an interaction between any two individuals is equally likely. Our approach implements the non-dimensional form of the standard SIR model because we couple it to the dimensionless forms of cost functions to find solutions to the social distancing problems. We start by providing a derivation of the non-dimensional SIR form starting from its standard form.

The differential equations describing the dynamics of the susceptible, infectious, and recovered individuals in the population are:

$$\begin{aligned} \frac{d}{dt}S &= -\frac{\beta SI}{N} \\ \frac{d}{dt}I &= \frac{\beta SI}{N} - \gamma I \\ \frac{d}{dt}R &= \gamma I \end{aligned} \tag{1}$$

where $S$ is the number of the susceptible, $I$ is the number of the infectious, $R$ is the number of the recovered, and $N$ is the total number of individuals in the population. The parameters $\beta$ and $\gamma$ respectively represent the transmission and recovery rate and have units of inverse time.

##### 1.1.1 Non-Dimensionalization

As is typically done, we define the dimensionless unit of time $\tau$, which is:

$$\tau = \gamma t. \tag{2}$$

In other words, in the dimensionless SIR model, $\tau = 1$ represents the average duration an individual is infectious for. We further recast the state variables in their dimensionless form where

$$\begin{aligned} s &= S/N, \\ i &= I/N, \\ r &= R/N. \end{aligned} \tag{3}$$

The dimensionless differential equations for the population dynamics become:

$$\begin{aligned} \frac{d}{d\tau}s &= -\frac{\beta}{\gamma}si, \\ \frac{d}{d\tau}i &= \frac{\beta}{\gamma}si - i, \\ \frac{d}{d\tau}r &= i. \end{aligned} \tag{4}$$

By defining the reproduction number $R_0 = \frac{\beta}{\gamma}$, these equations can be formulated as

$$\begin{aligned}\frac{d}{d\tau}s &= -R_0 si,\\ \frac{d}{d\tau}i &= R_0 si - i,\\ \frac{d}{d\tau}r &= i.\end{aligned} \tag{5}$$

##### 1.1.2 Final Epidemic Size

Without mitigation measures, and under the assumption that the initial conditions for the proportion of the population infected is very small (i.e., $i(\tau = 0) \ll 1$), and nearly the entire population is initially susceptible (i.e., $s(\tau = 0) \approx 1$), the proportion of the population that remains susceptible as $\tau \to \infty$ is given by the solution to the equation [Diekmann(2013)]

$$s_\infty = e^{-R_0(1-s_\infty)}, \tag{6}$$

and the final epidemic size is $1 - s_\infty$.

##### 1.1.3 The Effective Reproduction Number and Herd Immunity

The effective reproduction number $R_\tau$ describes the average number of infections produced by each infected individual in the population at time $\tau$. In the standard SIR model without mitigation measures,

$$R_\tau = sR_0. \tag{7}$$

Herd immunity occurs when the number of those susceptible in the population is reduced either by natural infection or vaccination such that the effective reproduction number is less than 1. Therefore, the maximum proportion of susceptible individuals in a population with herd immunity is

$$s_H = \frac{1}{R_0}. \tag{8}$$

#### 1.2 Time-Dependent Transmission

In classic SIR models, $\beta$ and $R_0$ are constant parameters. This paper considers the case where the transmission rate, $\beta$, is dynamic and influenced by behavioral changes. We will use the notation that $\beta$ is a dynamic variable and $\beta_0$ is the transmission rate without any behavioral mitigation. Similarly, we define $R_D(\tau)$ as the dynamic reproduction number, which is a function of social distancing behavior. $R_0$ is the reproduction number in the absence of any behavioral mitigation. In a dynamic framework, the dimensionless SIR model, Equations 5 become:

$$\begin{aligned}\frac{d}{d\tau}s &= -R_D si,\\ \frac{d}{d\tau}i &= R_D si - i,\\ \frac{d}{d\tau}r &= i.\end{aligned} \tag{9}$$

In the framework with a dynamic transmission rate, the effective reproduction number becomes:

$$R_\tau = sR_D. \tag{10}$$

##### 1.3 Cost Functions

We want to find the social distancing policy that minimizes a total cost, which includes the cost of social distancing and the cost of infections over a pre-specified time horizon ranging from time $t = 0$ to $t = t_{\text{final}}$. The social distancing policy is described by $\beta(t)$, the transmission rate, that depends on social distancing behavior at time $t$. We further define $\beta_0$ to be the infection rate for the disease in the absence of any behavioral change. Therefore, $\beta(t)$ is restricted to the interval $(0, \beta_0]$ The total cost is:

$$\text{Total cost} = (\text{Social distancing cost}) + (\text{Cost of infections}) \tag{11}$$

$$\int_{t=0}^{t_{\text{final}}} H_{\text{total}}(t)dt = \int_{t=0}^{t_{\text{final}}} \left[H_{\text{sd}}(t) + H_{\text{infect}}(t)\right] dt, \tag{12}$$

where $H_{\text{sd}}(t)$ is the cost of social distancing per unit time, and $H_{\text{infect}}$ is the cost of infections. As the transmission rate $\beta(t)$ decreases, the cost of social distancing per unit time, $H_{\text{sd}}(t)$, increases, but the cost of infections per unit time $H_{\text{infect}}(t)$ decreases.

###### 1.3.1 Cost of Infections

The number of new infections per unit time is $\frac{\beta SI}{N}$, which comes from the SIR equations. If we define $D$ to be the average cost of infection per infected individual. Then, the cost per unit time of infections is:

$$H_{\text{infect}}(t) = D\frac{\beta(t)S(t)I(t)}{N}. \tag{13}$$

Note that the total cost of infections is:

$$\text{Cost of infections} = D[R(t_{\text{final}}) + I(t_{\text{final}})]. \tag{14}$$

###### 1.3.2 Cost of social distancing

We assume that the total cost of social distancing is proportional to the size of the population $N$, and a cost parameter $C$, which has units cost per person per unit time. Therefore, we define the cost per unit time of social distancing to be:

$$H_{\text{sd}}(t) = NCg(\beta(t)/\beta_0). \tag{15}$$

Here, $g(x)$ is the function that defines the relationship between the relative cost of social distancing and the relative reduction in the transmission parameter. The function $g(x)$ should have the following properties:

1. **$\mathbf{g(x)}$ is a monotonically decreasing function on the interval $[0, 1]$.** The theoretical setup of the problem determines this property. The cost of social distancing should increase as the transmission rate is further decreased.

2. $\lim_{\mathbf{x}\to\mathbf{0}^+} \mathbf{g(x)} = \infty$. In theoretical terms, it is not possible to stop all transmission completely; therefore, the cost of decreasing transmission rates to zero should be infinite. From a practical perspective, this restriction will prevent optimal solutions to $\beta(t)$ from passing through $\beta(t) = 0$, and if the initial condition for $\beta(t)$ is positive, the solution will remain positive.

3. $\frac{\mathbf{d}}{\mathbf{dx}}\mathbf{g(x)}|_{\mathbf{x=1}} = \mathbf{0}$. Theoretically, if there is no cost of infections, the optimal $\beta(t) = \beta_0$. Therefore, the cost of social distancing should be minimized when $\frac{\beta(t)}{\beta_0} = 1$. From a practical perspective, combined with condition 2, this condition ensures that solutions to $\beta(t)$ will be bounded to the interval $(0, \beta_0]$ if correctly initialized to the interval.

4. **g(1)=0**. When $\frac{\beta(t)}{\beta_0} = 1$, there are no behavioral changes so the cost of social distancing should be zero.

Based on these three required properties, we choose the function to have the form

$$g(x) = -\ln(x) + x - 1, \tag{16}$$

and thus, substituting this into equation 15, the cost per unit time of social distancing is

$$H_{\text{sd}}(t) = -NC \ln\left(\frac{\beta}{\beta_0}\right) + NC\frac{\beta}{\beta_0} - NC. \tag{17}$$

###### 1.3.3 Dimensionless Cost Equations

We now derive the dimensionless form of the cost equation. We define a dimensionless cost of social distancing, $c$ to be:

$$c = \frac{C}{D\gamma}. \tag{18}$$

Effectively, $c$ compares the cost of social distancing to the cost of infection. Recall that $C$ has dimensions of cost per individual per unit time, $D$ has dimensions of cost per individual, and $\gamma$ has units of inverse time. Therefore, $c$ is dimensionless. When $c$ is small, people will more willingly social-distance as they feel that the cost of becoming ill outweighs the benefits of mixing socially. Their preference favors their lives and health compared to livelihoods and non-health-related well-being. When $c$ is large, people's preferences are switched. We define the dimensionless cost:

$$h = \frac{H}{DN}. \tag{19}$$

We can make the following substitutions in all equations:

$$\begin{aligned} \frac{\beta_0}{\gamma} &= R_0, \\ \frac{\beta}{\gamma} &= R_D. \end{aligned} \tag{20}$$

Then, the dimensionless cost of infection, Equation 13, becomes:

$$h_{\text{infect}} = R_D si. \tag{21}$$

The dimensionless cost of social distancing, Equation 15, becomes:

$$h_{\text{SD}} = cg(R_D/R_0). \tag{22}$$

We denote the cost function as $h(s, i, R_D)$ which can be expressed as:

$$h(s, i, R_D) = cg(R_D/R_0) + R_D si. \tag{23}$$

By setting $\mathcal{T}_{\text{final}} = \gamma T_{\text{final}}$, the total cost in its dimensionless form is

$$\text{Total Cost} = \int_0^{\mathcal{T}_{\text{final}}} \left[cg(R_D/R_0) + R_D si\right] d\tau. \tag{24}$$

##### 1.4 Myopic Solution

Here, we derive a myopic solution for $R_D$, where we assume that individuals or policymakers select the $R_D$ that minimizes the cost in Equation 23 at every moment in time. In other words

$$\frac{d}{dR_D}h(s, i, R_D) = 0. \tag{25}$$

This condition becomes:

$$-c\frac{R_0}{R_D} + \frac{c}{R_0} + si = 0 \tag{26}$$

solving for $R_D$, we have:

$$R_D = \frac{cR_0}{c + R_0 si} \tag{27}$$

#### 2 Paradoxical Case

In Figure 1 of the manuscript, the case in which $c = 0.05$ and $R_0 = 3$ has an apparent paradox: the dynamic reproduction number $R_D$ is always lower in the myopic case compared to the full optimization, however the total number of infected individuals is slightly higher by the end of the time frame in the myopic case. The full optimization reached herd immunity at about $\tau = 20$ and the myopic solution reaches herd immunity just before $\tau = 40$. We see that there are almost no new infections (and therefore almost no currently infected individuals) in the full optimization when herd immunity is achieved, and a small number of new


###### Acknowledgements

We wish to thank the National Cancer Institute (R21CA157571), and the National Institute of Allergies and Infectious Diseases (R01AI118705 & R01AI160240) for providing support in projects that led to preliminary work and ideas that motivated this project. Dr. Nowak acknowledges support from the Blodwen S. Huber Early Career Green and Gold Professor in Pathology and Laboratory Medicine at The Robert Larner, M.D. College of Medicine as well as support from The National Institute of General Medical Sciences 3P20GM125498-04S1.[rv]We thank Mr. Peng Dai and Dr. Sze-chuan Suen at the University of Southern California, for helpful discussions and on an earlier collaboration on related work. [Dai et al.(2021)Dai, Vardavas, Nowak, and Suen]


###### Disclaimer

The results and conclusions drawn on this working paper have not been peer-reviewed and do not necessarily represent the opinions of the RAND Corporation.

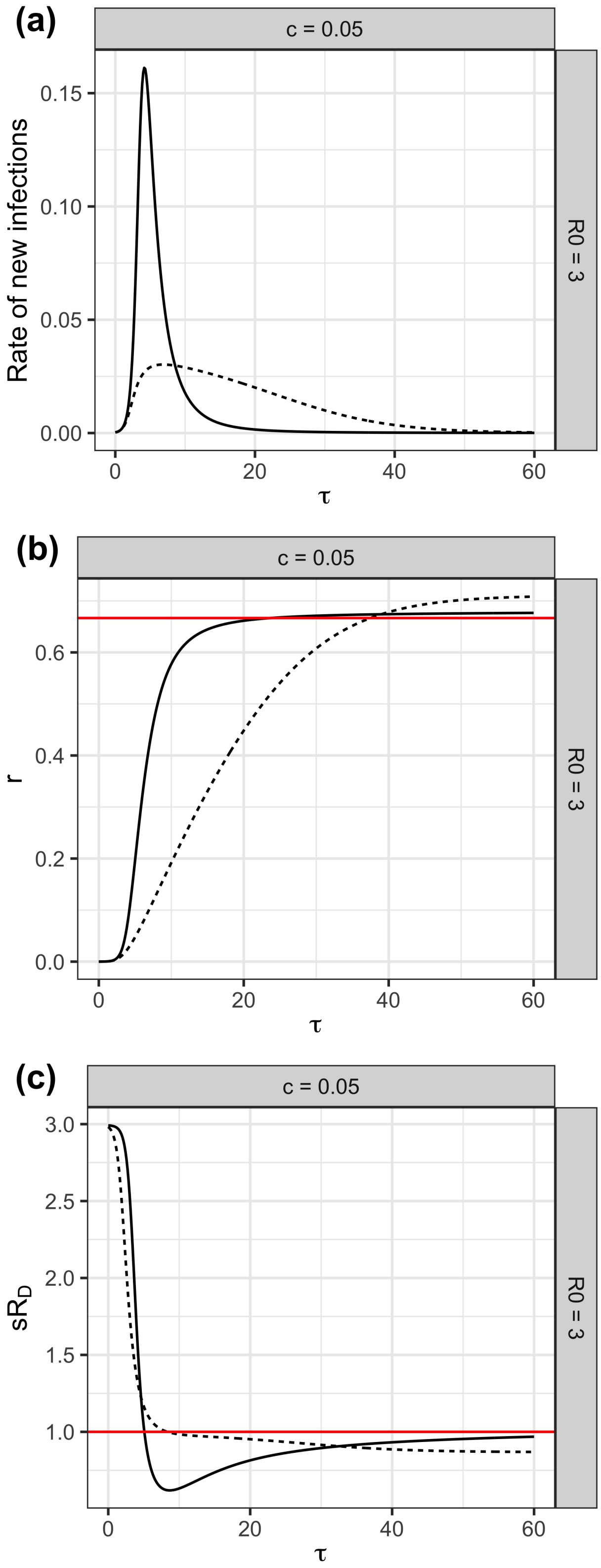


Figure 1: This figure shows additional results for the case in which $c = 0.05$ and $R_0 = 3$ from Figure 1 in the main text. Full optimization results are shown with the solid line and myopic results are shown with the dashed line. Panel (a) shows the rate of new infections. Panel (b) shows the number of recovered individuals over time where the red line shows the minimum number needed for herd immunity to be reached, and (c) shows the effective reproduction number, $sR_D$.
